# GDNF levels regulate lumbar motor neuron physiology and determine life expectancy in limb-onset ALS

**DOI:** 10.64898/2025.12.05.25341683

**Authors:** Soophie Olfat, Peyman Choopanian, Kärt Mätlik, Mehdi Mirzaie, Jaan-Olle Andressoo

## Abstract

Approximately one-third of amyotrophic lateral sclerosis (ALS) patients die within 12 months, while up to 10% live more than a decade. The factors driving this variation remain unclear but may elucidate disease mechanisms and guide therapy development. Here, we analyzed raw postmortem RNA-seq datasets from ALS patients and controls to quantify glial cell line-derived neurotrophic factor (GDNF) expression across cervical (n=161), thoracic (n=44), and lumbar (n=143) spinal cord regions. GDNF levels varied several-fold between individuals and correlated with motor neuron marker gene expression specifically in the lumbar spinal cord, but not in thoracic or cervical regions. In 76 limb-onset ALS patients, higher lumbar GDNF expression correlated with longer survival, whereas no such relationship was observed in 32 bulbar-onset patients. To test causality, we used the GDNF hypermorph SOD1^G93A^ mouse model, in which ∼30% (skeletal muscle) and 2-fold (spinal cord) GDNF overexpression enhanced neuromuscular junction (NMJ) preservation and improved motor function for 10–13 weeks, equating to ∼8–10 years in human life expectancy. In contrast, previous ectopic GDNF applications yielded only transient benefit. These findings identify endogenous lumbar GDNF levels as a determinant of life expectancy in limb-onset ALS and suggest that strategies to stimulate GDNF signalling may slow disease progression, with potentially the greatest benefit in leg-onset patients.

## Introduction

Life expectancy after ALS diagnosis varies dramatically, ranging from less than one year to over a decade [1]. Although ALS is invariably progressive, often leading to severe disability within 1–2 years, the biological factors underlying this more than ten-fold variation remain unknown. Identifying such determinants could provide mechanistic insight into disease pathogenesis, enable subtype stratification, and guide development of tailored therapies.

Glial cell line-derived neurotrophic factor (GDNF) is a potent survival factor for motor neurons (MNs). Deletion of genes encoding GDNF or its receptors GFRα1 and RET causes ∼25% reduction in MN numbers, particularly in the lumbar spinal cord of mice [2], [3]. Conversely, GDNF overexpression in muscle increases lumbar MN numbers by ∼10% [4]. In ALS models, ectopic GDNF delivery via various systems supports MN survival and delays symptom onset by up to 3 weeks [4]–[7] (Supplementary Table 1). These preclinical findings motivated a first-in-human phase 1/2a trial, where neural progenitor cells overexpressing GDNF were transplanted into the lumbar spinal cord of 18 ALS patients [8]. Although the procedure was generally safe, efficacy data have not yet been reported.

Despite extensive work in animal models, it remains unclear how endogenous GDNF levels vary in humans, whether such variation modulates MN physiology, and whether it influences ALS progression. To address these questions, we re-analyzed publicly available RNA-seq data from healthy individuals and ALS patients across spinal cord regions. We then tested causality in a GDNF-hypermorph SOD1^G93A^ mouse model, where endogenous GDNF levels are increased post-transcriptionally [9] at a range comparable to that observed in human samples.

## Material and methods

### Analysis of human samples

#### Muscle

Publicly available bulk-RNASeq profiles were obtained from the European Genome Archive, identified as Dataset ID EGAS00001005904, and can be downloaded at ega-archive.org. This dataset consists of 128 samples gathered from 20 healthy individuals. Moreover, the muscle transcriptomics atlas is open for exploration via a ShinyApp, from which we obtained the normalized gene expression, original sites: profile. https://elifesciences.org/articles/80500 and https://tabbassidaloii.shinyapps.io/muscleAtlasShinyApp/.

#### Spinal cord

To investigate GDNF expression in ALS patients and controls, we used RNASeq from [10]. This data originates from postmortem samples from cervical (n=161), thoracic (n=44), and lumbar (n=143) spinal cord regions. The normalized gene expression profiles can be downloaded from https://zenodo.org/records/6385747. Analysis was performed using R, and the code will be available upon request.

#### Animals

All animal experiments were conducted in accordance with the 3R principles of the EU directive 2010/63/EU governing the care and use of experimental animals and authorized by the County Administrative Board of Southern Finland (license numbers ESAVI/9280/2018 and ESAVI/34289/2022). All protocols were authorized by the National Animal Experiment Board of Finland. The mice were provided with *ad libitum* access to food and water at temperature-controlled conditions at 20 to 22 ºC under a 12h/12h light/dark cycle at a relative humidity of 50-60%. Bedding material and cages (aspen chips, Tapvei Oy, Finland) were changed weekly, along with wooden tubes and aspen shavings as enrichment. Male SOD1^G93A^ mice (strain B6SJL-TgN (SOD1^G93A^)1Gur-Low SOD1^G93A^ copy number) from The Jackson Laboratory (Bar Harbor) were used in this study. Transgene expression was assessed by PCR (tail DNA) and qPCR using the following primers: SOD1^G93A^ fwd: 5’-CAC GTG GGC TCC AGC ATT-3’; SOD1^G93A^ rev: 5’-TCA CCA GTC ATT TCT GCC TTT G-3’; internal CTR fwd: 5’-GGG AAG CTG TTG TCC CAA G-3’; internal CTR rev: 5’-CAA GGG GAG GTA AAA GAG AGC-3’. Generation and genotyping of GDNF hypermorphic (*Gdnf*^*wt/hyper*^, referred to as GDNFh) and Tg(SOD1^G93A^) transgenic mice have been previously described [9], [11]. Male *Gdnf*^*wt/hyper*^; SOD1^G93A^ and *Gdnf*^*wt/wt*^; SOD1^G93A^ littermates were used in the analysis.

#### Behavioral tests

Behavioral tests, including rotarod, coat hanger, and open field, were performed as described previously by Mätlik et al [12].

#### Humane endpoint

Animals were sacrificed if they exhibited weight loss of more than 15-20% compared to their weight before the onset of symptoms or based on the animal’s ability to right themselves 30 seconds after being placed on their side. All animals were monitored at least twice a week.

#### Tissue collection

For tissue collection from adult mice, animals were divided into 3 groups. Group 1: 8-week-old *Gdnf*^wt/hyper^ and WT animals to measure GDNF mRNA levels in muscle. Group 2: WT+ SOD1^G93A^ and *Gdnf*^wt/hyper^ + SOD1^G93A^ animals at the mid-stage of the disease (34-35 weeks). Group 3: WT+ SOD1^G93A^ and *Gdnf*^wt/hyper^ + SOD1^G93A^ animals at the end stage of the disease (at humane endpoint). Mice were anesthetized with sodium pentobarbital (100 mg/kg intraperitoneally [i.p.]) and intracardially perfused with PBS followed by 4% paraformaldehyde (PFA) in 0.1 M phosphate buffer (pH 7.4). The spinal cord and muscle were collected for further analysis. For tissue collection at E18.5, females were sacrificed, and muscle tissue from the hind limbs containing gracilis, rectus femoris, semitendinosus, and vastus muscles was isolated from the embryos and snap frozen for later analysis.

#### Genotyping

Genomic DNA was isolated from the tail using Extracta DNA Prep for PCR - tissue (cat. no. 95091, Quanta Biosciences, USA). Genotyping was performed using AccuStart II GelTrack PCR SuperMix (cat. no. 95136, Quanta Biosciences, USA). The following primers were used for SOD1^G93A^ tg mice: h SOD1^G93A^ forward, CAT CAG CCC TAA TCC ATC TGA; h SOD1^G93A^ reverse, CGC GAC TAA CAA TCA AAG TGA. PCR products were analyzed on a 1.5-2% agarose gel in borate buffer. *Gdnf*^wt/hyper^ mice were genotyped as described previously [9].

#### Analysis of protein levels

GDNF Emax Immunoassay System (G7620, Promega) was used to measure GDNF protein levels, according to the manufacturer’s protocol. Mouse muscle samples were homogenized in a lysis buffer recommended by the manufacturer, and the homogenate was centrifuged at 5000 rpm for 15 minutes at 4 °C. The supernatant was acid-treated using HCl, and GDNF protein levels and total protein levels were measured and analyzed as described previously in [13]. Samples from *Gdnf*^*cKO*^ x Nestin-Cre mice were used as a control [14].

#### Immunohistochemistry

Mice were anesthetized with pentobarbital (Mebunat, 200 mg/kg, i.p., Yliopiston Apteekki) and perfused with PBS and 4% formaldehyde (Sigma). Spinal cord samples were post-fixed in 4% formaldehyde for 24 hours and dehydrated in 30% sucrose (Thermo Fisher Scientific) in PBS before sectioning. Sucrose-dehydrated tissues were cryosectioned to a thickness of 40 µm. c-RET immunostaining was performed on serially cut lumbar spinal cord tissue sections (RET: Neuromics, Cat.no: GT15002; dilution 1:100). The secondary antibodies used were Alexa Fluor 568, Abcam, Cat.no: ab175704 (1:500). The samples were imaged with a Leica 122 Stellaris 8 FALCON/DLS confocal microscope and a Nikon Ti inverted microscope using a Yokagawa CSU-X1, 10,000 rpm spinning disk, a back-illuminated sCMOS camera (Photometrics Kinetix, 95% QE, 6,5 μm pixels), and the NIS-Elements software with a 20x objective. Quantification and analysis of motor neurons were performed by following a previously published protocol [15].

#### Assessment of the neural muscular junction

Gastrocnemius muscles were sectioned longitudinally into 20 µm slices using a cryostat. Muscle sections were blocked with PBS-Triton X-100 (0.5%) and incubated for 24h with anti-SV2 and anti-2H3 primary antibodies (1:100, DSHB, AB_2315387, and AB_531793), recognizing neurofilament (2H3) and synaptic vesicles (SV2) to visualize the axons and pre-synaptic nerve terminals. After washes, sections were incubated with Alexa488-conjugated α-bungarotoxin (α-BTX) to visualize the endplate (stains acetylcholine receptors in skeletal muscle fibers)(1:200, Life Technologies, B13422) and Alexa Fluor Plus 555 donkey anti-mouse secondary antibody (1:500, Thermo Fisher Scientific). Axons and NMJs were imaged using a Nikon Ti inverted microscope using a Yokagawa CSU-X1, 10,000 rpm spinning disk, a back-illuminated sCMOS camera (Photometrics Kinetix, 95% QE, 6,5 μm pixels), and the NIS-Elements software with a 40x objective. For quantitative analysis, NMJs were classified based on the degree of innervation of postsynaptic receptor plaques by nerve terminals. About 20 endplates were analyzed in each animal. The number of endplates of each category was presented as a percentage of the total number of counted NMJs.

### Data analysis

As outlined above, publicly available bulk RNA-seq datasets were used to investigate the natural variation in GDNF expression among control human samples and its association with motor neuron (MN) genes. Data from human spinal cord and muscle samples were investigated to assess this variation. Data from the human spinal cord and muscles were analyzed to investigate this variation. Since the normalized data had already been processed and published, reanalysis of raw data to extract expression levels was avoided. Instead, normalized expression values were employed to visualize and explore specific genes of interest, including performing correlation analyses for selected targets. These analyses were performed in R (version 4.4.2), utilizing ggplot2 for data visualization and ggpubr for calculating and plotting correlation values.

For data generated in this study, including behavioral tests, immunohistochemistry, and neuromuscular junction analysis, statistical tests and visualizations were conducted using GraphPad Prism (version 8.4.2, GraphPad Software, USA). Behavioral performance metrics (e.g., rotarod, coat hanger, open field) were analyzed using one-way ANOVA and post hoc comparisons where appropriate, while neuromuscular junction data were classified and quantified based on the degree of innervation. All statistical results are presented as mean ± SEM, with significance defined as P < 0.05.

## Results

In mice, Gdnf mRNA levels correlate linearly with GDNF protein across CNS and peripheral tissues [9], [13], [16], supporting the use of RNA-seq as a proxy for protein abundance.

Analysis of leg muscle RNA-seq from healthy individuals (128 samples gathered from 20 healthy individuals) showed ∼3-fold inter-individual variation in GDNF expression (Fig. 1). In spinal cord, variation reached ∼6-fold, with GDNF correlating strongly with MN marker expression in the lumbar but not cervical or thoracic regions (cervical; n=161, thoracic; n=44, and lumbar; n=143 spinal cord regions.)(Fig. 2). These findings align with mouse genetics, where GDNF or RET deletion causes selective lumbar MN loss by up to 25% [2], [3], while developmental GDNF overexpression increases lumbar MN numbers by 5-10% [4]. Together, these observations suggest that endogenous GDNF influences MN physiology in a region-specific manner and may modulate disease outcome in limb-onset ALS.

**Figure 1.**
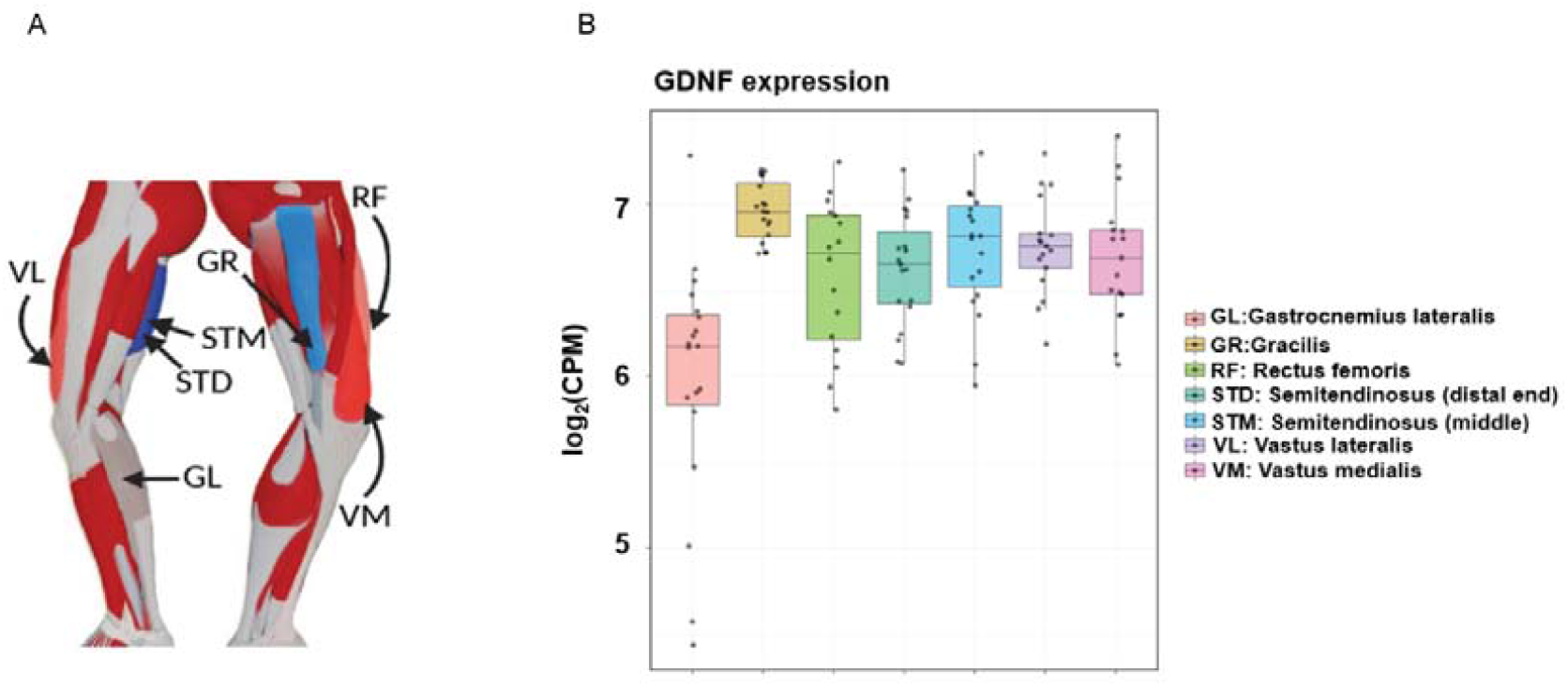
Analysis of natural variation of GDNF mRNA expression levels in 7 skeletal muscles from 20 healthy individuals, raw data acquired from [27]. **(A)** A schematic overview of the leg muscles. Arrows point to the muscles that were included in this study, and the abbreviations are opened on B. **(B)** The distribution of GDNF mRNA expression levels across seven different muscles. Each point in the boxplots represents an individual sample. GDNF mRNA levels vary about 3-fold between healthy individuals.

**Figure 2.**
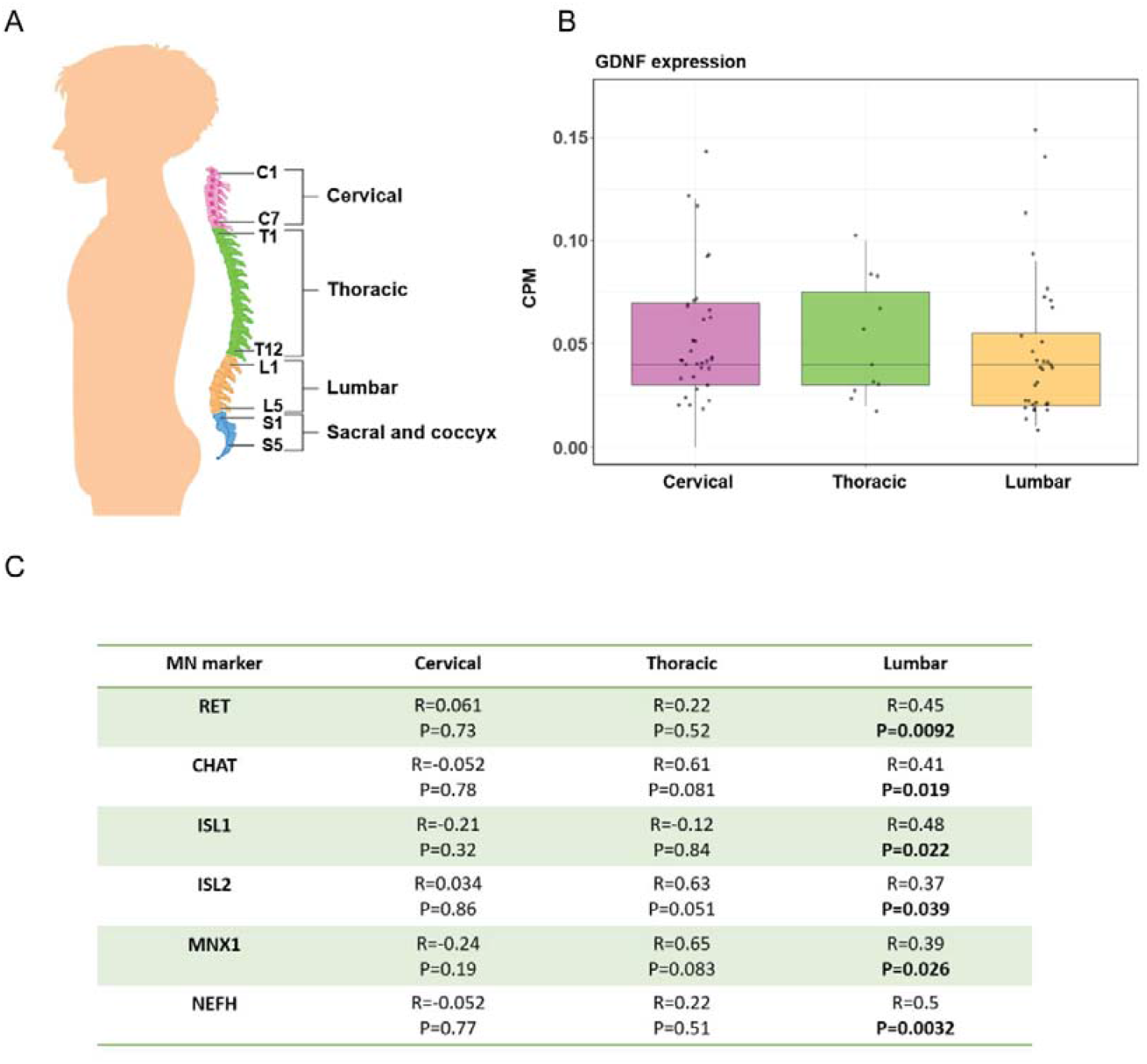
Analysis of natural variation of GDNF mRNA expression levels in cervical (n=161), thoracic (n=44), and lumbar (n=143) spinal cord and correlation analysis with MN marker gene expression levels (A) A schematic overview of the spinal cord with cervical, thoracic, and lumbar segments indicated. **(B)** Analysis of raw RNA-SEQ data downloaded from [10] the human spinal cord shows that GDNF mRNA levels vary up to about 6-fold between healthy individuals. **(C)** Correlation analysis shows a positive correlation between GDNF mRNA expression and MN marker gene mRNA expression levels in the lumbar but not cervical or thoracic spinal cord; significant P-values are highlighted in bold.

To address this, we analyzed spinal cord RNA-seq from ALS patients. Higher lumbar GDNF levels correlated with significantly longer survival in limb-onset cases (n = 76), but not in bulbar-onset patients (n = 32) (Fig. 3). Data distinguishing upper versus lower limb onset were unavailable.

**Figure 3.**
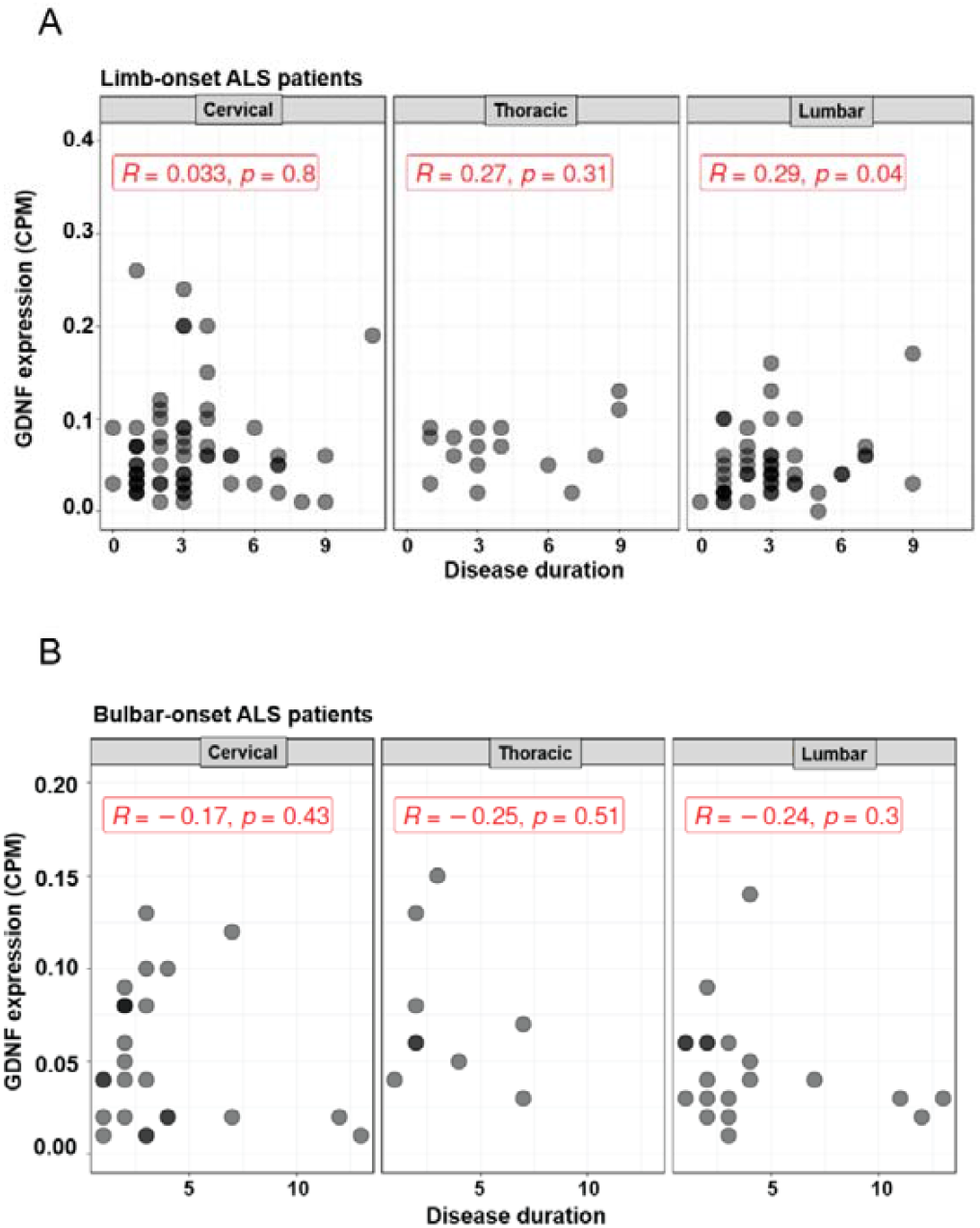
Correlation analysis of GDNF expression levels in cervical (n=161), thoracic (n=44), and lumbar (n=143) regions of the spinal cord with disease duration (life-expectancy) in ALS patients (A) Limb onset ALS patients (76 limb-onset ALS patients) **(B)** Bulbar onset ALS patients (32 bulbar-onset ALS patients). The correlation coefficients and P-values are displayed and highlighted in red. A significant positive correlation is observed between GDNF expression levels in the lumbar spinal cord and disease duration in Limb-onset ALS (P-value=0.04).

We next tested causality using Gdnf hypermorphic (*Gdnf*^*hyper*^)mice (Fig. 4A), where 3′UTR replacement increases endogenous expression by ∼2-fold without ectopic overexpression [9], [13], which associates with side-effects [17]–[23]. In muscle, GDNF protein was elevated by ∼20–30% in heterozygotes and ∼50% in homozygotes at E18.5, with similar increases in mRNA at 2 months (Fig. 4B–C), consistent with earlier spinal cord data [9].

**Figure 4.**
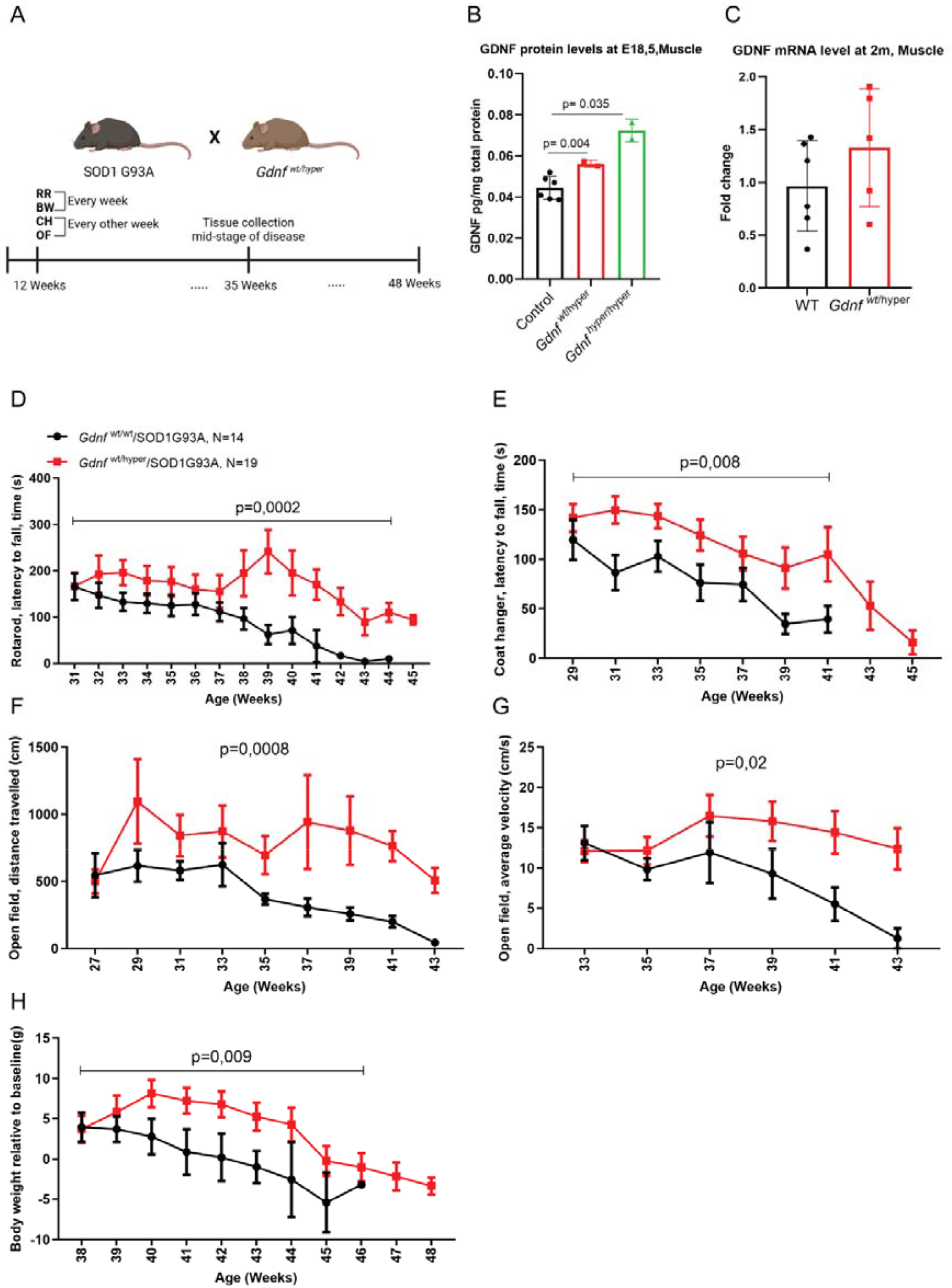
Analysis of endogenous GDNF levels effects on limb-onset ALS progression using GDNF Hypermorph allele in SOD1^G93A^ limb-onset ALS model. **(A)** Schematic of experimental plan, SOD1^G93A^ mice were crossed to *Gdnf*^wt/hyper^ mice, behavioral tests were started at 12 weeks of age, Accelerating RotaRod test (RR) and Body Weight (BW) were measured every week, and Open Field (OF) and Coat Hanger (CH) tests were performed every other week. At week 35, when the genotypes started to show significant differences in behavioral tests, the tissues in a subset of animals were collected for analysis. **(B)** GDNF protein levels were measured with an ELISA test at E18.5 in hind limb skeletal muscle tissue and showed a *Gdnf*^hyper^ allele dose-dependent increase (t-test, p=0.004 for controls vs *Gdnf*^wt/hyper^ and p=0.035 for controls vs *Gdnf*^hyper/hyper^). **(C)** GDNF mRNA levels measurement in 2-month-old (2m) mice in the hind limb skeletal muscle. Longitudinal analysis of male *Gdnf*^wt/wt^ and *Gdnf*^wt/hyper^ mice. **(D)** Accelerating Rotarod (RR) test, **(E)** Coat Hanger test (CH), **(F)** Open Field (OF) test, distance traveled **(G)**, OF velocity analysis **(H)**, body weight (BW) measurement, p-values indicate statistical difference between the genotypes.

Crossing these mice with SOD1^G93A^ (Fig. 4A) revealed improved motor performance from ∼week 31 onward, including rotarod, coat hanger, and open field tests, with benefits persisting for 10–13 weeks (Fig. 4D–G and S Fig. 1A-D). ALS end-stage-associated weight loss was also ameliorated (Fig. 4H and S Fig. 1E).

At week 35, when genotypes were already split for the most endpoints, MN soma counts (RET+) were similar between genotypes (Fig. 5A). However, NMJs were markedly preserved in *Gdnf*^*wt/hyper*^; SOD1^G93A^ mice (>80% innervated endplates vs. ∼60% in controls) (Fig. 5B–C). Thus, modest physiological upregulation of GDNF delays ALS progression by preserving NMJ integrity rather than preventing MN loss.

**Figure 5.**
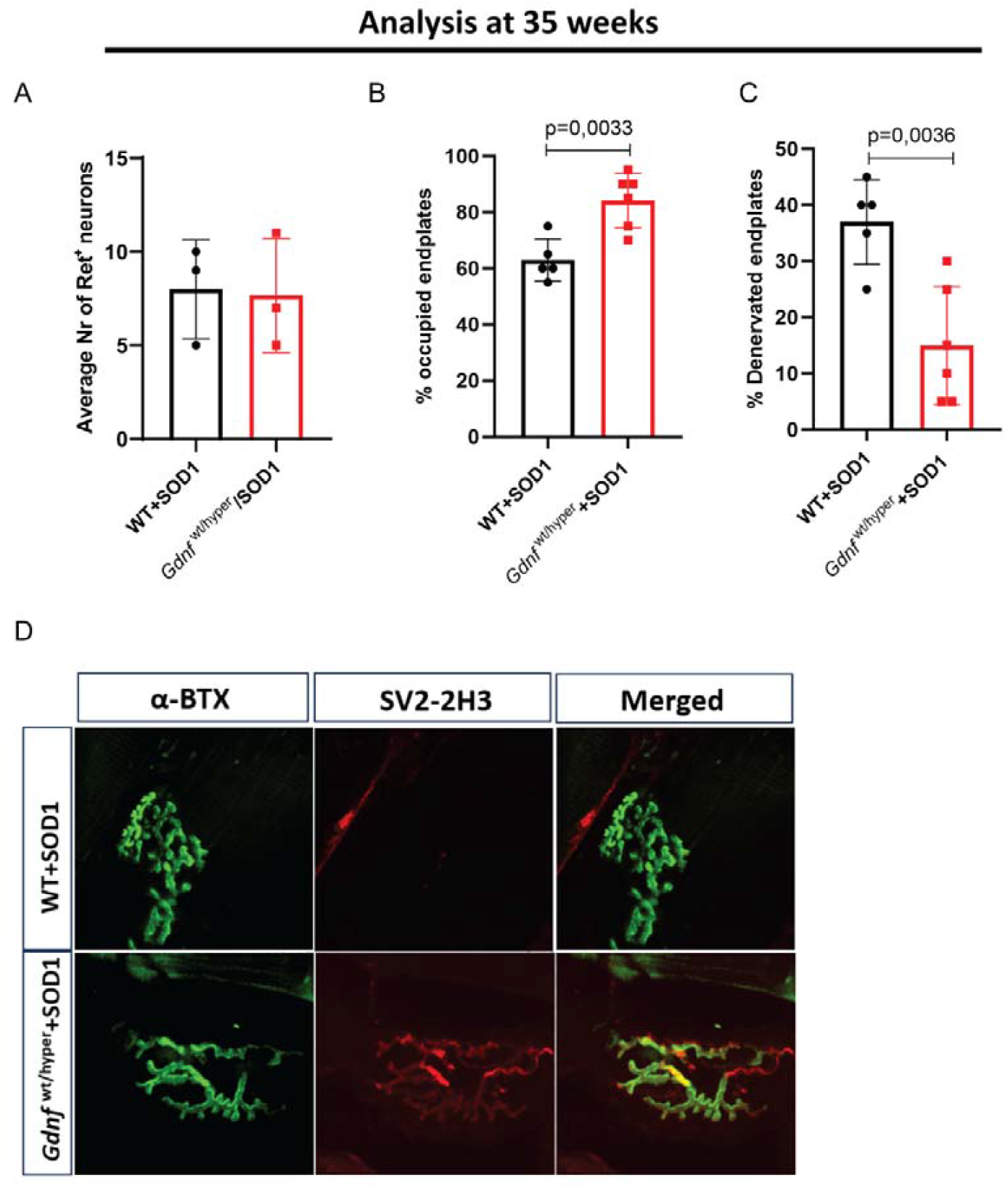
Analysis of motor neuron (MN) and neuromuscular junction (NMJ) number at week 35. **(A)** Analysis of RET-positive neurons in the lumbar spinal cord (n=3 animals per group, 6 sections per animal) reveals no difference between the genotypes **(B-C)** Analyses of NMJs at 35 weeks of age showed significantly more occupied (p=0.0033) NMJs in *Gdnf*^wt/hyper^ mice and less denervated (p=0.0036) NMJs compared to the *Gdnf*^wt/wt^ animals (n=5 neurofilament (2H3) and synaptic vesicles (SV2) were stained to visualize the axons and pre-synaptic nerve terminals, α-bungarotoxin (α-BTX) was used to visualize the endplate (stains acetylcholine receptors in skeletal muscle fibers). (*Gdnf*^*wt/wt*^ n=5 and n=6 *Gdnf*^wt/hyper^). **(D)** Representative images of an NMJ in a 35-week-old *Gdnf*^wt/wt^ and *Gdnf*^wt/hyper^ mouse in SOD1^G93A^ background (scale bar = 10µm). Note the lack of neuronal staining in *Gdnf*^wt/wt^ in SOD1^G93A^ background, indicating degeneration of MN axons.

## Discussion

In mice, the role of GDNF in motor neuron (MN) development is well established. Deletion of *Gdnf, Gfra1*, or *Ret* during embryogenesis or before postnatal day 5 reduces lumbar MN numbers by ∼20–25%, whereas deletion after this period has no effect on MN survival or neuromuscular junctions (NMJs) [3]–[7]. Conversely, developmental GDNF overexpression in skeletal muscle increases lumbar MN numbers by 5–10% [4]. Based on these findings, ectopic GDNF delivery has been tested in ALS models, but the outcomes vary: improvements in motor function generally last only 0–3 weeks, likely reflecting differences in site, level, and timing of expression [4]–[7] (Supplementary Table 1). For example, developmental transgenic muscle-specific GDNF overexpression extended motor function in SOD1^G93A^ mice for ∼15 days, whereas astrocyte-specific overexpression had no effect [6].

Our data suggest that lumbar MN dependence on GDNF signalling is conserved in humans. GDNF mRNA levels correlated with expression of multiple MN markers in the lumbar but not cervical or thoracic spinal cord. Moreover, lumbar GDNF levels correlated with disease duration in limb-onset ALS, indicating that endogenous GDNF preferentially modulates progression in lower-limb–onset ALS. Direct comparison with upper-versus lower-limb onset awaits further clinical stratification.

To test whether modest increases in endogenous GDNF can affect ALS outcome, we used the *Gdnf*^*hyper*^ allele, which elevates GDNF levels by ∼30% in muscle and ∼2-fold in spinal cord. In SOD1^G93A^ mice, this genetic background prolonged motor function by ∼12–13 weeks compared to littermate controls. This benefit is markedly greater than in prior ectopic delivery studies (0–3 weeks) and, when scaled to human lifespan, could correspond to nearly a decade of additional preserved function. The improvement was not due to enhanced baseline performance (Supplementary Figure 1) but reflected superior preservation of NMJs, although not MN soma, at symptomatic stages.

Future work should assess whether increasing endogenous GDNF after symptom onset, for example, using tamoxifen-inducible Cre and conditional hypermorph alleles, can also confer benefit. Testing small-molecule GDNF pathway agonists currently in development for Parkinson’s disease [24]–[26] in limb-onset ALS models is of particular translational interest. Finally, establishing links between tissue GDNF levels (e.g., muscle) and accessible fluids (CSF, serum) could help define GDNF as a biomarker for prognosis and stratification in ALS subgroups.

## Data Availability

All data produced in the present study are available upon reasonable request to the authors

## Figure legends

**Supplementary figure 1.**
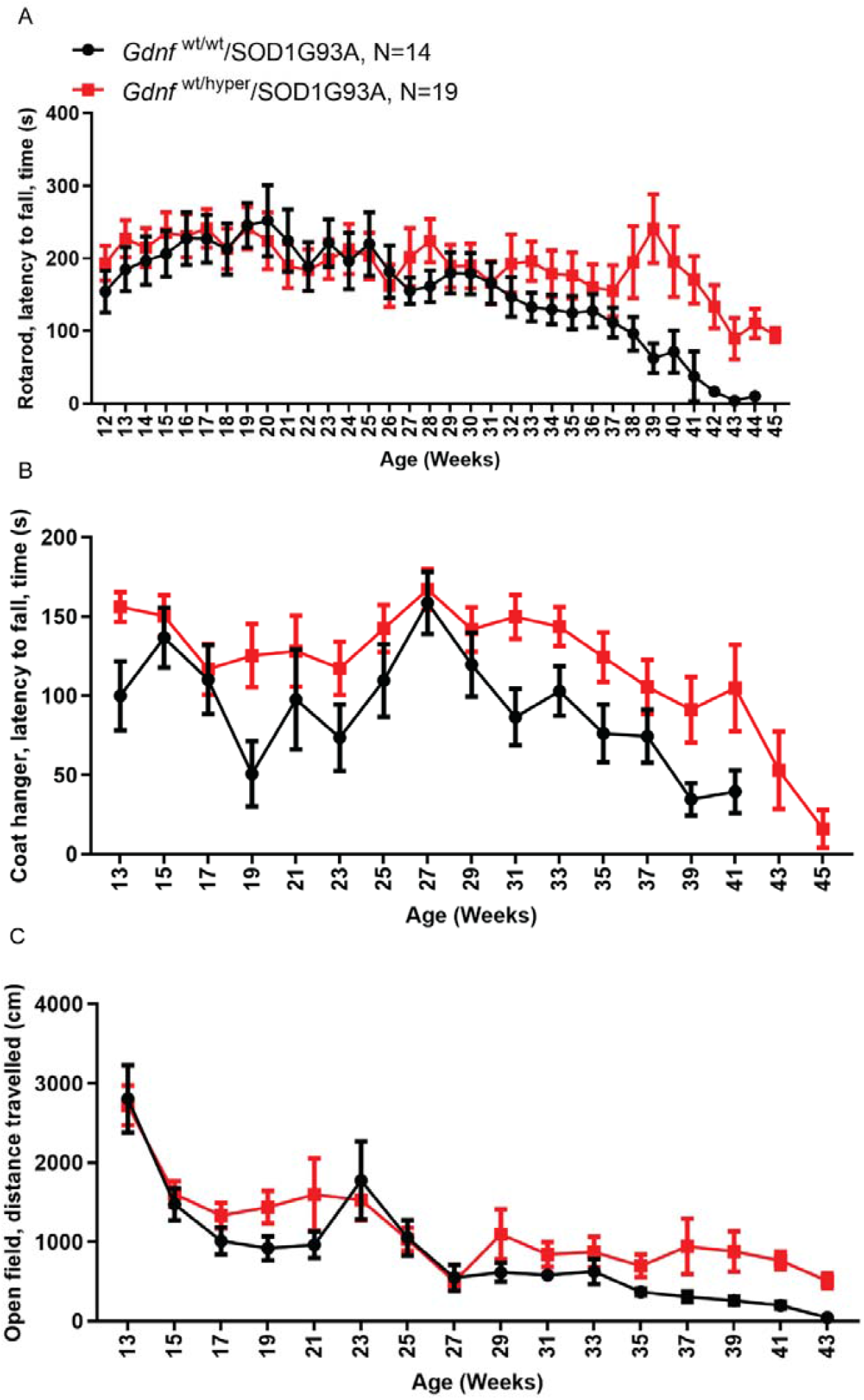
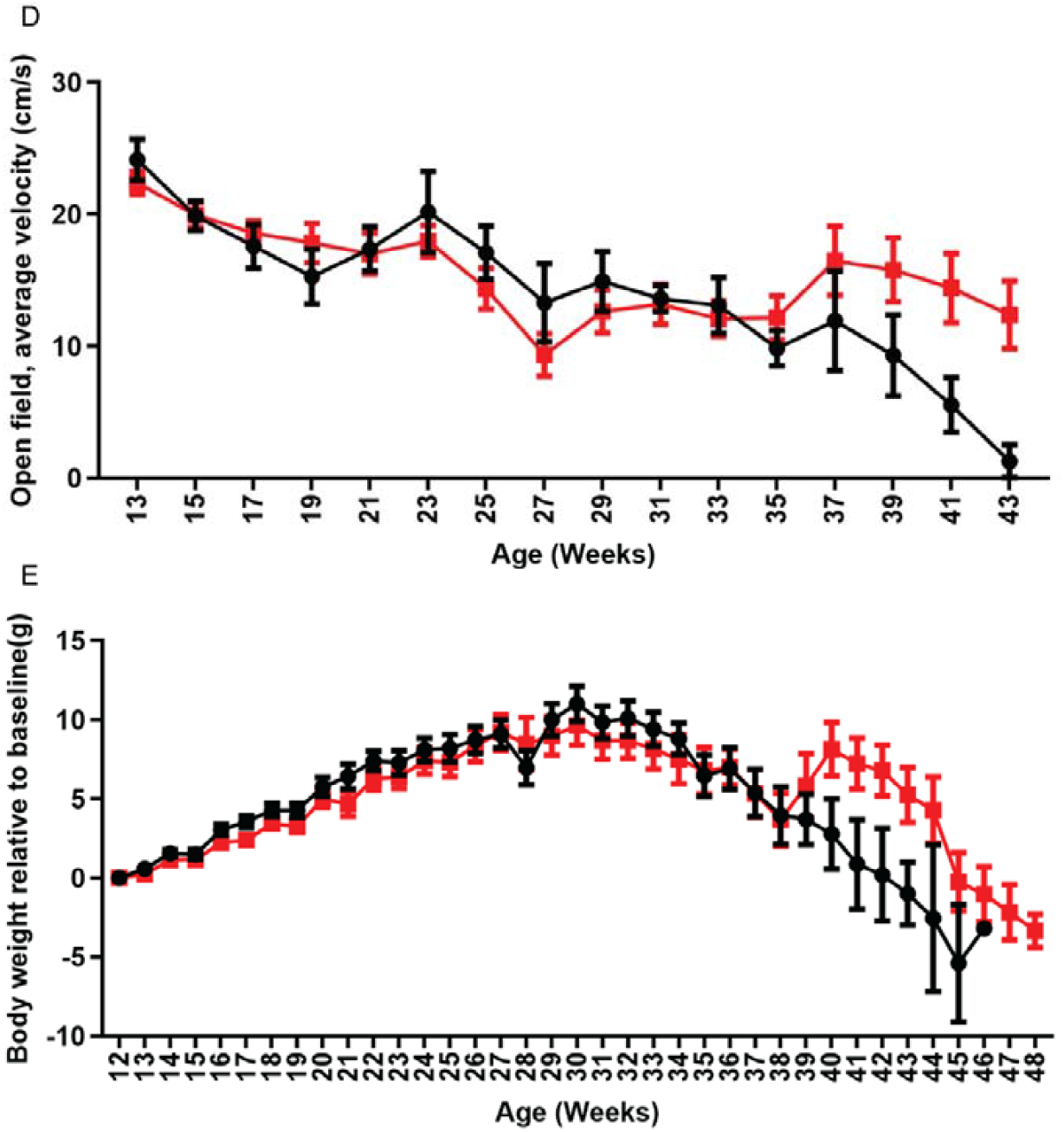
Analysis of endogenous GDNF levels effects on limb-onset ALS progression during the whole experimental period using GDNF Hypermorph allele (*Gdnf*^hyper^) in SOD1^G93A^ limb-onset ALS model. Male mice were longitudinally tested with the indicated motor tests **(A-D)** and for body weight **(E). (A)** Accelerating the rotarod test measures latency to fall in motor coordination and strength tasks. **(B)** The coat hanger test measures primarily muscle strength. **(C)** The open field test measures total distance traveled over 30 minutes. **(D)** The open field test average velocity reflects movement speed. **(E)** Body weight relative to the baseline.

## Acknowledgments

J.O.A. was supported by the Academy of Finland (grants no. 297727 and 350678), Sigrid Juselius Foundation, ERA-NET NEURON grant nr 352077, Center of Innovative Medicine (CIMED), Hjärnfonden, Swedish Research Council (grants no. 2019-01578 and 2022-01093), Helsinki Institute of Life Science, European Research Council (ERC, grant no. 724922), Åhlens Foundation, JAES Foundation grant no. 240034 and HORIZON-RIA grant nr 101188432 DTRIP4H.Kärt Mätlik was supported by a fellowship from the Alfred Kordelin Foundation. This study was performed at the Live Cell Imaging Core facility/Nikon Center of Excellence, at Karolinska Institutet, Sweden, supported by the KI infrastructure council. The BioImage Informatics Facility at SciLifeLab, National Microscopy Infrastructure (VRRFI 2019-00217), and recipient of the Chan-Zuckerberg Initiative, assisted in the image analysis. The authors thank Jenni Lahtinen for her technical help.

---

**Supplementary Table 1.**
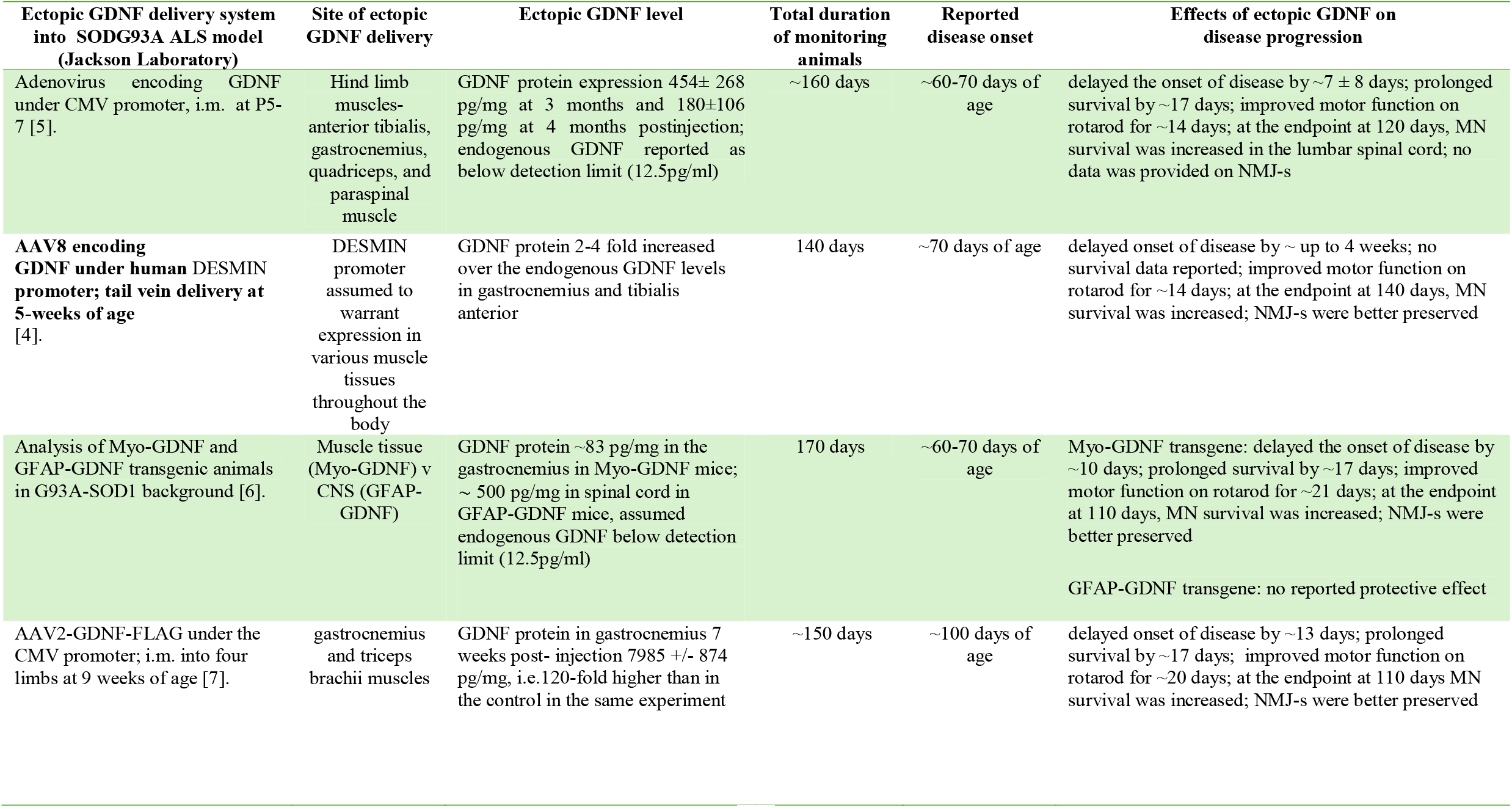
Ectopic GDNF deliveries that have been tested in SOD1^G93A^ rodent ALS model. Abbreviations: i.m. - intramuscular delivery via injection; P-post-natal day.

| Ectopic GDNF delivery system into SODG93A ALS model (Jackson Laboratory) | Site of ectopic GDNF delivery | Ectopic GDNF level | Total duration of monitoring animals | Reported disease onset | Effects of ectopic GDNF on disease progression |
| --- | --- | --- | --- | --- | --- |
| Adenovirus encoding GDNF under CMV promoter, i.m. at P5-7 [5]. | Hind limb muscles- anterior tibialis, gastrocnemius, quadriceps, and paraspinal muscle | GDNF protein expression $454 \pm 268$ pg/mg at 3 months and $180 \pm 106$ pg/mg at 4 months postinjection; endogenous GDNF reported as below detection limit (12.5pg/ml) | ~160 days | ~60-70 days of age | delayed the onset of disease by $\sim 7 \pm 8$ days; prolonged survival by $\sim 17$ days; improved motor function on rotarod for $\sim 14$ days; at the endpoint at 120 days, MN survival was increased in the lumbar spinal cord; no data was provided on NMJ-s |
| **AAV8 encoding GDNF under human DESMIN promoter; tail vein delivery at 5-weeks of age** [4]. | DESMIN promoter assumed to warrant expression in various muscle tissues throughout the body | GDNF protein 2-4 fold increased over the endogenous GDNF levels in gastrocnemius and tibialis anterior | 140 days | ~70 days of age | delayed onset of disease by ~ up to 4 weeks; no survival data reported; improved motor function on rotarod for $\sim 14$ days; at the endpoint at 140 days, MN survival was increased; NMJ-s were better preserved |
| Analysis of Myo-GDNF and GFAP-GDNF transgenic animals in G93A-SOD1 background [6]. | Muscle tissue (Myo-GDNF) v CNS (GFAP-GDNF) | GDNF protein $\sim 83$ pg/mg in the gastrocnemius in Myo-GDNF mice; $\sim 500$ pg/mg in spinal cord in GFAP-GDNF mice, assumed endogenous GDNF below detection limit (12.5pg/ml) | 170 days | ~60-70 days of age | Myo-GDNF transgene: delayed the onset of disease by $\sim 10$ days; prolonged survival by $\sim 17$ days; improved motor function on rotarod for $\sim 21$ days; at the endpoint at 110 days, MN survival was increased; NMJ-s were better preserved |
|  |  |  |  |  | GFAP-GDNF transgene: no reported protective effect |
| AAV2-GDNF-FLAG under the CMV promoter; i.m. into four limbs at 9 weeks of age [7]. | gastrocnemius and triceps brachii muscles | GDNF protein in gastrocnemius 7 weeks post- injection $7985 \pm 874$ pg/mg, i.e. 120-fold higher than in the control in the same experiment | ~150 days | ~100 days of age | delayed onset of disease by $\sim 13$ days; prolonged survival by $\sim 17$ days; improved motor function on rotarod for $\sim 20$ days; at the endpoint at 110 days MN survival was increased; NMJ-s were better preserved |

